# Impact of mass testing during an epidemic rebound of SARS-CoV-2: A modelling study

**DOI:** 10.1101/2020.12.08.20246009

**Authors:** Paolo Bosetti, Cécile Tran Kiem, Yazdan Yazdanpanah, Arnaud Fontanet, Bruno Lina, Vittoria Colizza, Simon Cauchemez

**Affiliations:** Mathematical Modelling of Infectious Diseases Unit, Institut Pasteur, UMR2000, CNRS, Paris, France; Collège Doctoral, Sorbonne Université, Paris, France; Infections Antimicrobials Modelling Evolution (IAME) UMR 1137, University of Paris, Paris, France; Emerging Diseases Epidemiology Unit, Institut Pasteur, Paris, France; PACRI Unit, Conservatoire National des Arts et Métiers, Paris, France; National Reference Center for Respiratory Viruses, Department of Virology, Infective Agents Institute, North Hospital Network, Lyon, France; Virpath Laboratory, International Center of Research in Infectiology, INSERM U1111, CNRS—UMR 5308, École Normale Supérieure de Lyon, Université Claude Bernard Lyon, Lyon University, Lyon, France; INSERM, Sorbonne Université, Pierre Louis Institute of Epidemiology and Public Health, Paris 75012, France

## Abstract

We used a mathematical model to evaluate the impact of mass testing in the control of SARS-CoV-2. Under optimistic assumptions, we find that one round of mass testing may reduce daily infections by up to 20-30%. As a consequence, very frequent testing would be required to control a quickly growing epidemic if other control measures were to be relaxed. Mass testing is most relevant when epidemic growth remains limited, thanks to a combination of interventions.

## Introduction

Several European countries facing a large increase in COVID-19 cases have moved back into lockdown. While Test-Trace-Isolate remains for now the most efficient way to control an epidemic rebound at the end of these lockdowns, there is debate about optimal ways to use testing (1, 2). So far, testing has mostly targeted symptomatic individuals and contacts of cases. The increasing availability of diagnostic tests now makes it possible to consider a strategy of mass testing, that consists in testing a large proportion of the population in a single campaign to identify and isolate as many infected individuals as possible. The development of rapid antigenic tests facilitates the implementation of such an approach since these tests can provide results in less than 30 minutes compared to 1-2 days for the standard PCR. Although these antigenic tests have a reduced sensitivity as compared to the PCR test for the diagnostic of SARS-Cov-2, the most sensitive rapid antigenic tests have a sensitivity threshold that is sufficient to identify a large proportion of infectious individuals (with high viral shedding)(3).

However, even with antigenic tests, the implementation of mass testing will be challenging with an impact that is still to be determined. Here, using a mathematical model, we assess the possible impact of mass testing campaigns in a scenario of epidemic rebound at the end of the ongoing strong restrictions in Metropolitan France.

### Modelling the spread of SARS-CoV-2 and consecutive campaigns of mass testing

We use a compartmental SEIIR model to describe the spread of SARS-CoV-2 in Metropolitan France (4, 5). After infection, individuals move between the following compartments: compartment E1 where they have been exposed but are not infectious yet (average duration: 4 days), compartment E2 where they are infectious (i.e. can transmit) but have no symptoms (average duration: 1 day), compartment I where they are infectious and may be symptomatic (average duration 3 days), compartment R where they recovered. This description leads to a generation time of about 7 days consistent with existing data on chains of transmission and viral excretion (6, 7).

In our baseline scenario, we deliberately consider optimistic assumptions to derive an upper bound of the impact of mass testing. We assume that an infectious individual (i.e. in compartments E2 or I) has a probability of testing positive with an antigenic test equal to sensitivity Se=90% and that a person testing positive will reduce onward transmissions by ρ=70% on average thanks to isolation (1, 2). In sensitivity analyses, we also consider Se=60%, 75% and ρ=50%, 90%.

Assuming a basic reproduction number R0=0.9 from 30th November 2020, the start of the ongoing lockdown, to January 4th 2021, we expect about 8000 daily infections (with about half of them being detected) and 13% of the population of Metropolitan France being infected by January 4th 2021. From January 4th 2021, we consider rebound scenarios where the number of daily infections doubles every 10 (effective reproduction number Re=1.6), 14 (Re=1.4), 17 (Re=1.3) and 21 (Re=1.2) days. As a reference, Re was estimated at 1.4 from hospitalisation data in mid-October.

We assume that the first testing campaign starts on January 4th, is repeated every 1-30 days with 0-90% of the population being tested during each campaign.

### Impact of mass testing on epidemic dynamics

Figure 1 shows epidemic dynamics for monthly and bi-weekly campaigns of mass testing starting on January 4th 2021. For an epidemic rebound with a doubling time of 21 days, we would expect 140,000 daily infections by May 1st in the absence of mass testing. A monthly campaign of mass testing in which 75% of the population is being tested would reduce that number to 80,000 daily infections, and a bi-weekly campaign to 35000. For a doubling time of 14 days, a monthly campaign testing 75% of the population would reduce daily infections at the peak from 360,000 to 285,000 and bi-weekly campaigns to 200,000 daily infections.

**Figure 1.**
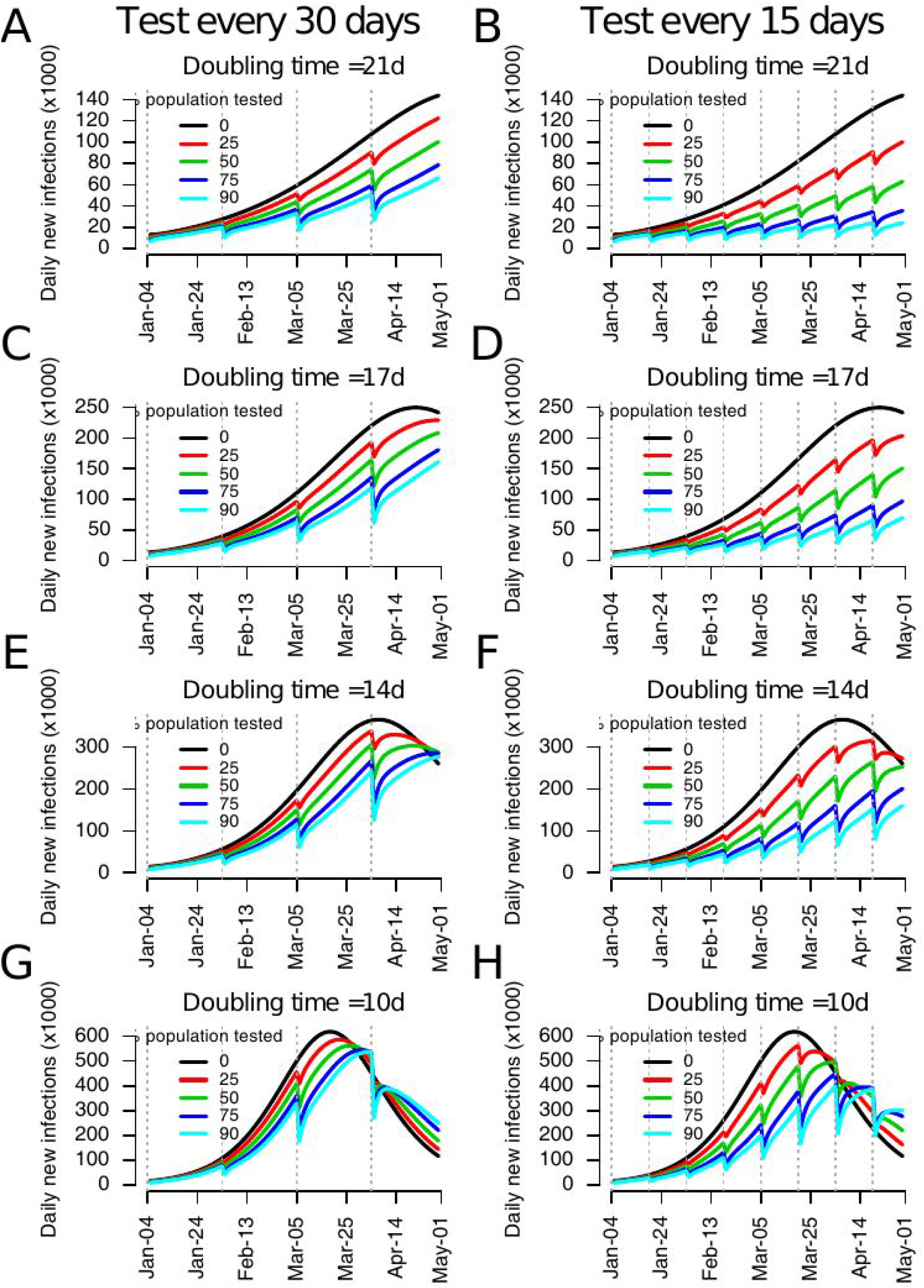
Expected numbers of daily infections when monthly (A, C, E, G) and bi-weekly (B, D, F, H) testing campaigns are performed targeting different proportions of the population. We explore epidemiological scenarios where the number of infections double every 21 (A, B), 17 (C, D), 14 (E, F) and 10 (G, H) days. We assume that the probability to detect an infectious individual with an antigenic test is Se=90% and the reduction in onward transmission following a positive test is ρ =70%.

In our baseline scenario, we find that a single campaign targeting 75% of the population reduces the number of infections that occur 10 days after the campaign by about 21% (Figure 2A). We obtain 14% and 18% reductions for a sensitivity Se of 60% and 75%, respectively, and 15% and 27% reductions for an effectiveness of isolation ρ of 50% and 90%, respectively. Results are insensitive to the value of the effective reproduction number (results not shown). These reductions may have limited impact on overall dynamics in a context of quick epidemic rebound. For example, in our baseline scenario, if the number of infections doubles every 21 days and 75% of the population is being tested, we expect that it will take 10 days to get back to the epidemiological situation observed prior to testing (Figure 2B). For a doubling time of 14 days, we only gain 6 days.

**Figure 2.**
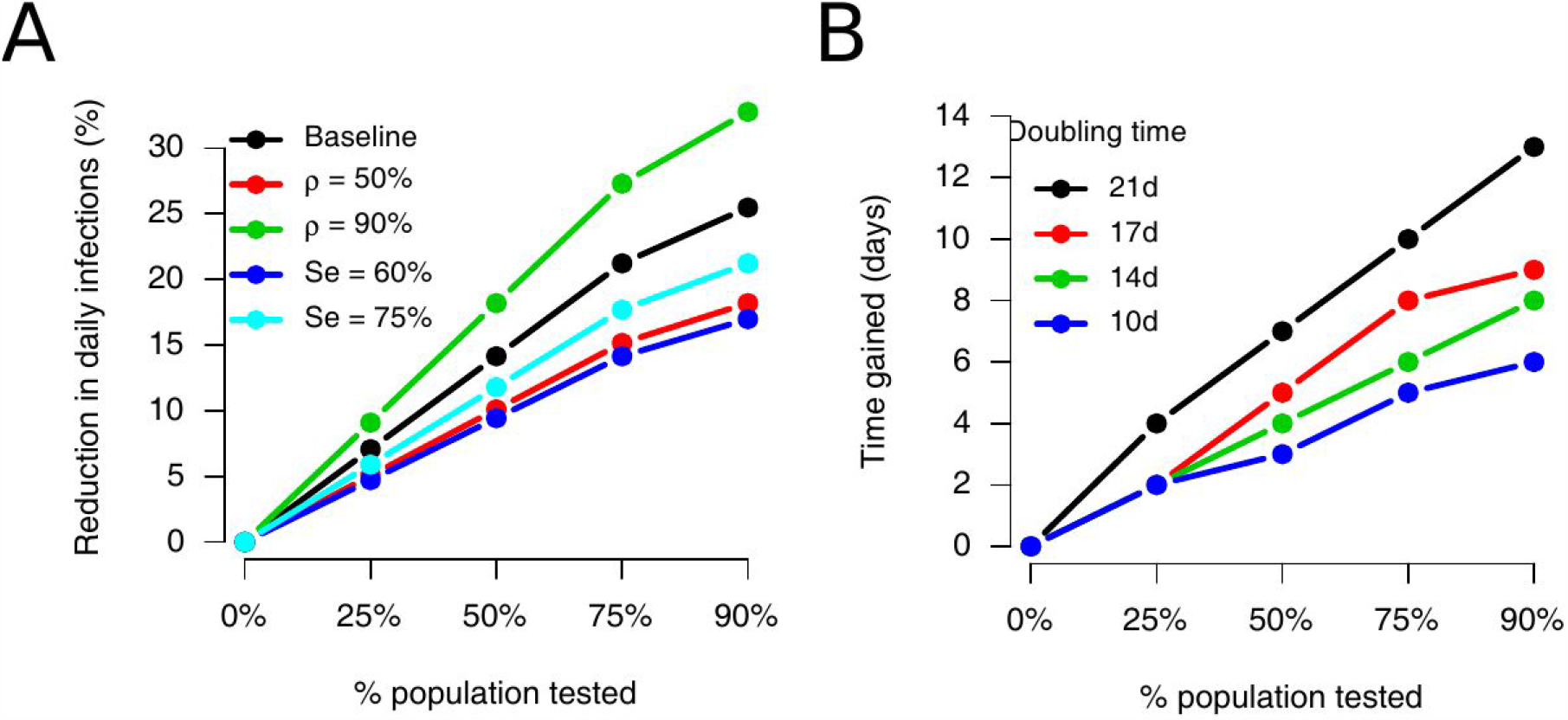
Impact of a single campaign of mass testing. A. Reduction in the number of infections occurring 10 days after the campaign, for different scenarios. In our baseline scenario (black line), the probability to detect an infectious individual with an antigenic test is Se=90% and the reduction in onward transmission following a positive test is ρ =70%. We consider alternative scenarios with ρ =50% (red line), ρ =90% (green line), Se=60% (blue line) and Se=75% (light blue line). B. Time it takes for the number of daily infections to go back to levels observed prior to mass testing, as a function of the doubling time in the baseline scenario.

Figure 3 shows the maximum daily number of infections observed by May 1st as a function of the number of days between consecutive campaigns and the proportion of the population being tested in each campaign. In our baseline scenario, for a doubling time of 21 days, to ensure that the number of infections remains below 80,000, 60,000 and 40,000 up to May 1st, a campaign testing 75% of the population would need to occur every 30, 22, and 16 days, respectively. For a doubling time of 14 days, testing of 75% of the population should occur every 6 days to have <40,000 daily infections.

**Figure 3.**
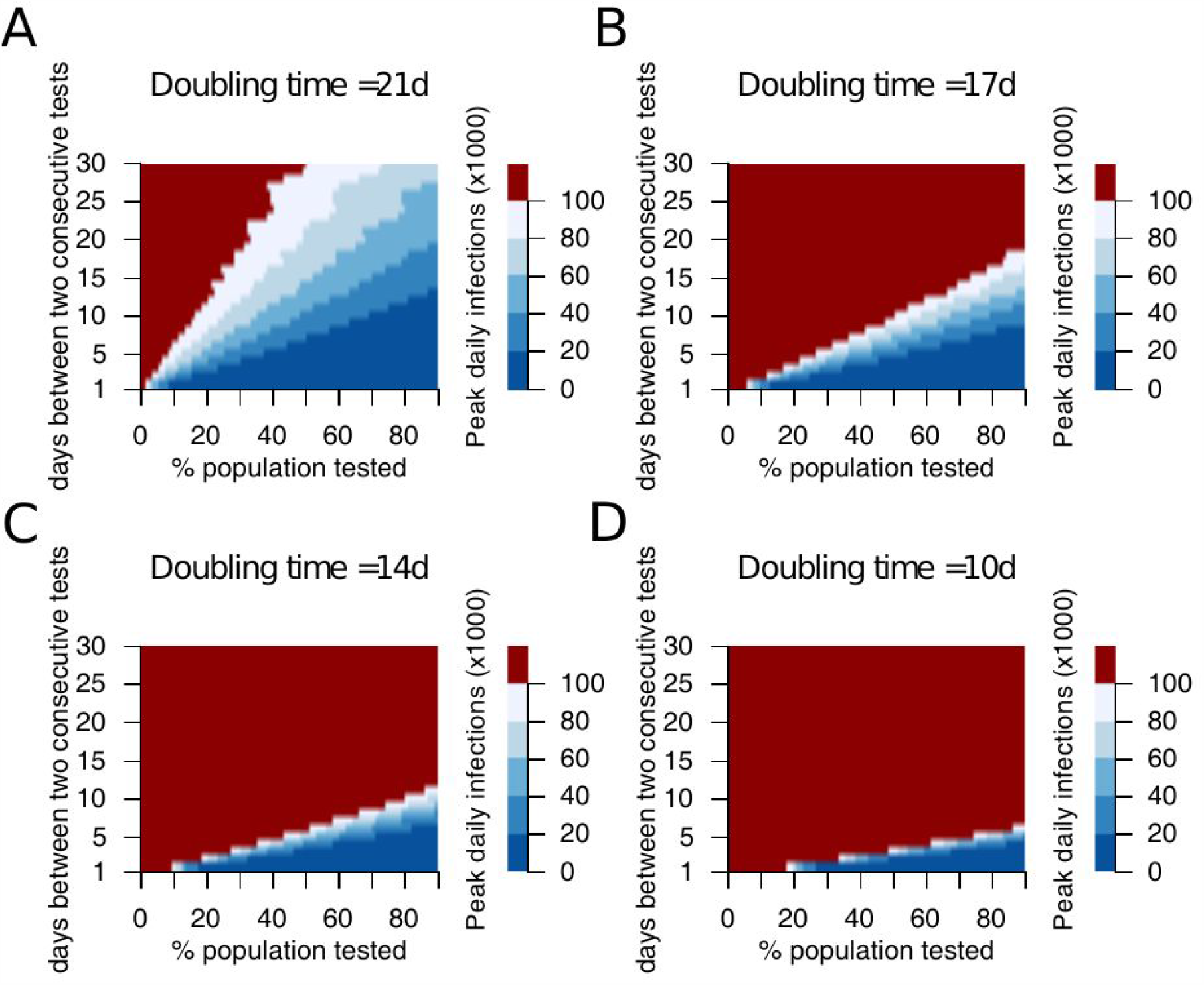
Expected maximum number of daily infections observed by May 1st as a function of the number of days between consecutive campaigns and the proportion of the population being tested in each campaign, for doubling times of 21 (A), 17 (B), 14 (C) and 10 (D) days. We assume that the probability to detect an infectious individual with an antigenic test is Se=90% and the reduction in onward transmission following a positive test is ρ =70%.

We explore the sensitivity of our results to model assumptions (Figure 4). To observe <40,000 daily infections by May 1st with a doubling time of 21 days, testing 75% of the population needs to be repeated every 21 days and 11 days when the effectiveness of isolation following a positive test is ρ =90% and ρ =50%, respectively. Such campaigns need to be repeated every 12 and 10 days for a sensitivity Se of the test of 75% and 60%.

**Figure 4.**
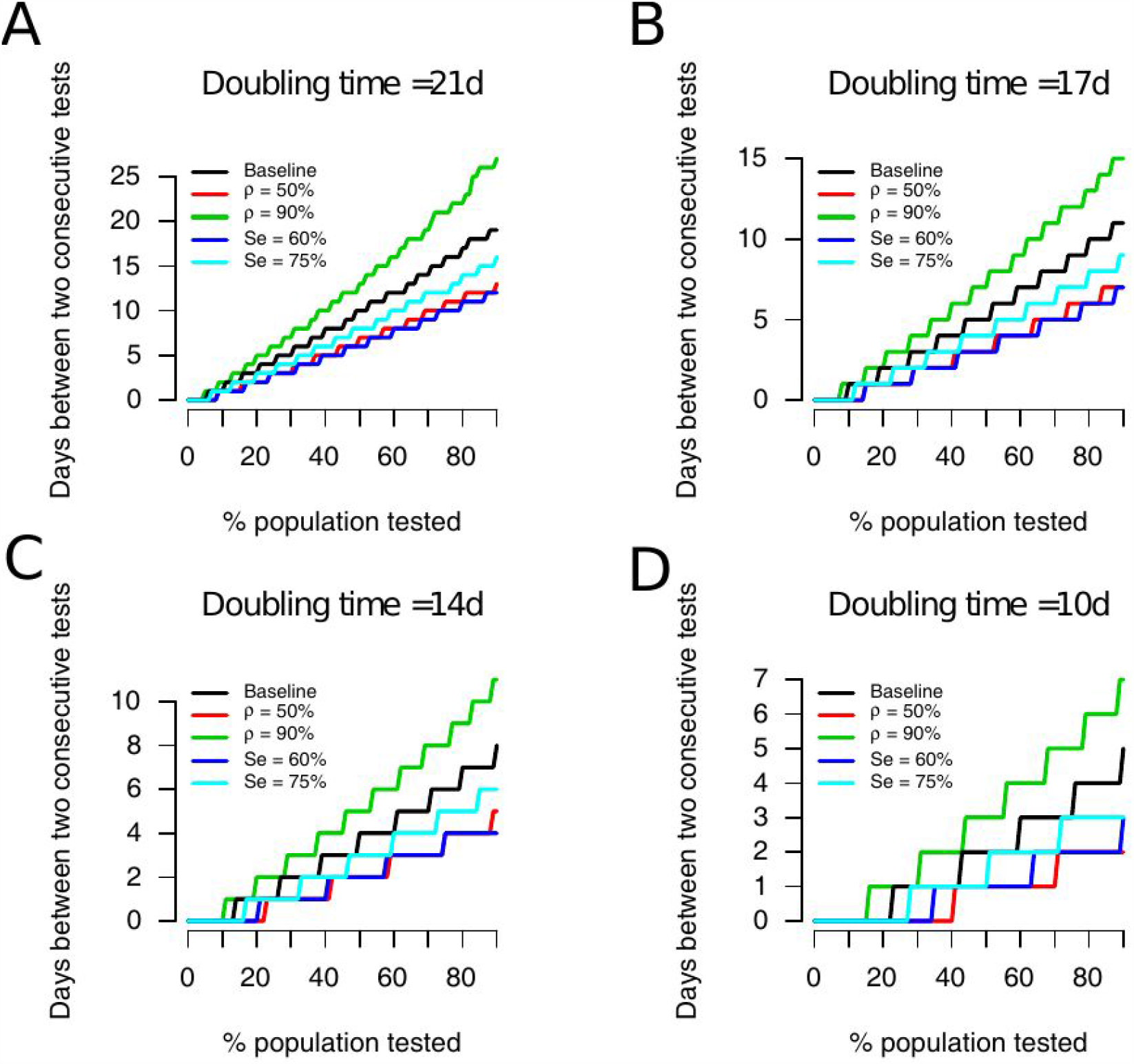
Frequency of mass testing campaigns necessary to keep the number of daily infections below 40,000 until May 1st as a function of the proportion of the population being tested in each campaign, for different modeling assumptions. In our baseline scenario (black line), we assume that the probability to detect an infectious individual with an antigenic test is Se=90% and the reduction in onward transmission following a positive test is ρ =70%. We consider alternative scenarios where the reduction in transmission is ρ =50% (red line) and 90% (green line) and where the probability to detect an infectious individual is Se = 60% (blue line) and Se = 75% (light blue line).

## Discussion

We used a simple mathematical model to highlight the potential and limits of mass testing for the control of SARS-CoV-2 epidemics. Under optimistic assumptions, we find that one round of mass testing may reduce daily infections by up to 20-30%. As a consequence, very frequent testing would be required to control a quickly growing epidemic if other control measures were to be relaxed. Mass testing is therefore most relevant when epidemic growth remains limited, thanks to a combination of interventions.These results are consistent with another modelling study (8).

In combination with interventions able to slow down the epidemic growth, high frequency of screening remains the key element to provide an additional contribution to epidemic control (7). However, the logistics, efficiency of isolation, and voluntary participation of the population to repeated mass screening need to be carefully assessed. Few pilot studies have been conducted so far in small regions and with one-shot approaches requiring several days of implementation (9). Limited adhesion to isolation if positive would largely compromise such effort. Here we considered an optimistic scenario with 70% reduction of onward transmission in isolation (with 50%-90% for sensitivity), but survey data from the UK point to smaller adhesion rates, of about 10%(10).

During a campaign of mass testing in Slovakia that occured in multiple rounds, the prevalence of infection dropped by about 60% in the week between two rounds (11). The campaign happened along with important restrictions, including a 1 week lockdown. This makes it difficult to dissociate the impact of mass testing from that of other interventions. For example, if the effective reproduction number was 0.6-0.7 thanks to other measures, we would expect the prevalence of infection to drop by about 30%-40% per week in the absence of mass testing. In such a scenario, mass testing could have contributed to the additional 20%-30% reduction that was observed, which would be roughly consistent with estimates under our most optimistic scenarios. In our assessment, we only considered strategies in which only cases are isolated. In contrast, in Slovakia, the whole household was quarantined when a case was detected, potentially leading to higher impact on spread.

An important limitation of mass testing is that, when the campaign begins, approximately half of the individuals who are infected are still in the latent phase E1 so that they do not shed sufficient virus to test positive. These individuals will not be detected and will become infectious after the campaign, fueling once again the epidemic. This problem can be partly mitigated by extending isolation to household contacts, as in Slovakia, and performing robust contact tracing of cases(12). Finally, for a given number of tests, impact might be higher if tests are target towards areas, populations or places of higher incidence.

Our study has a number of limitations. First it relies on a deterministic model that may imperfectly capture epidemic dynamics when the daily number of infection becomes very low. In such situations, the epidemic may take longer to rebound than anticipated by the model. Second, we do not address the logistical challenges involved in regular mass screening and assume that individuals are tested in a single day.

## Data Availability

No data referred in the manuscript.

## References

1. J. van Beek, et al., From more testing to smart testing: data-guided SARS-CoV-2 testing choices https:/doi.org/10.1101/2020.10.13.20211524.

2. A. Alemany, et al., Analytical and Clinical Performance of the Panbio COVID-19 Antigen-Detecting Rapid Diagnostic Test https:/doi.org/10.1101/2020.10.30.20223198.

3. M. J. Mina, R. Parker, D. B. Larremore, Rethinking Covid-19 Test Sensitivity - A Strategy for Containment. N. Engl. J. Med. (2020) https:/doi.org/10.1056/NEJMp2025631.

4. H. Salje, et al., Estimating the burden of SARS-CoV-2 in France. Science 369, 208–211 (2020).

5. A. Andronico, et al., Evaluating the impact of curfews and other measures on SARS-CoV-2 transmission in French Guiana https:/doi.org/10.1101/2020.10.07.20208314.

6. J. Paireau, et al., Early chains of transmission of COVID-19 in France https:/doi.org/10.1101/2020.11.17.20232264.

7. D. B. Larremore, et al., Test sensitivity is secondary to frequency and turnaround time for COVID-19 screening. Science Advances, eabd5393 (2020).

8. M. C. J. Bootsma, et al., Regular universal screening for SARS-CoV-2 infection may not allow reopening of society after controlling a pandemic wave https:/doi.org/10.1101/2020.11.18.20233122.

9. E. Holt, Slovakia to test all adults for SARS-CoV-2. Lancet 396, 1386–1387 (2020).

10. L. E. Smith, et al., Adherence to the test, trace and isolate system: results from a time series of 21 nationally representative surveys in the UK (the COVID-19 Rapid Survey of Adherence to Interventions and Responses [CORSAIR] study) https:/doi.org/10.1101/2020.09.15.20191957.

11. M. Pavelka, et al., The effectiveness of population-wide, rapid antigen test based screening in reducing SARS-CoV-2 infection prevalence in Slovakia https:/doi.org/10.1101/2020.12.02.20240648.

12. J.A. M. López, et al., Anatomy of digital contact tracing: role of age, transmission setting, adoption and case detection https:/doi.org/10.1101/2020.07.22.20158352.

